# Safe and reliable transport of prediction models to new healthcare settings without the need to collect new labeled data

**DOI:** 10.1101/2023.12.13.23299899

**Authors:** Rudraksh Tuwani, Andrew Beam

## Abstract

How can practitioners and clinicians know if a prediction model trained at a different institution can be safely used on their patient population? There is a large body of evidence showing that small changes in the distribution of the covariates used by prediction models may cause them to fail when deployed to new settings. This specific kind of dataset shift, known as covariate shift, is a central challenge to implementing existing prediction models in new healthcare environments. One solution is to collect additional labels in the target population and then fine tune the prediction model to adapt it to the characteristics of the new healthcare setting, which is often referred to as localization. However, collecting new labels can be expensive and time-consuming. To address these issues, we recast the core problem of model transportation in terms of uncertainty quantification, which allows one to know when a model trained in one setting may be safely used in a new healthcare environment of interest. Using methods from conformal prediction, we show how to transport models safely between different settings in the presence of covariate shift, even when all one has access to are covariates from the new setting of interest (e.g. no new labels). Using this approach, the model returns a prediction set that quantifies its uncertainty and is guaranteed to contain the correct label with a user-specified probability (e.g. 90%), a property that is also known as coverage. We show that a weighted conformal inference procedure based on density ratio estimation between the source and target populations can produce prediction sets with the correct level of coverage on real-world data. This allows users to know if a model’s predictions can be trusted on their population without the need to collect new labeled data.

## Introduction

There has been increased adoption of machine learning models across various domains, including healthcare^1–3^. Though not universally true, these models promise improved predictive performance and efficiency gains over traditional statistical approaches in certain scenarios^2^, such as high-dimensional or non-linear prediction tasks. However, their successful deployment in sensitive application areas like healthcare remains challenging. One significant obstacle is that patient populations and care practices vary substantially between different hospitals and healthcare systems^4,5^, and strict data privacy protections and institutional constraints make the sharing of patient data across organizations difficult. This severely limits the ability to train robust machine learning models that generalize across diverse medical settings.

One solution to this problem is to collect new labeled data at each healthcare site and use it to adapt or retrain models to local conditions through a process called *localization* or *fine-tuning*^6^ . However, this can be prohibitively expensive and time-consuming depending on how difficult it is to create a labeled dataset in the target environment. Moreover, many popular machine learning model families like random forests do not lend themselves to simple fine-tuning using new data. This often necessitates training models from scratch for each new healthcare environment.

For reliable deployment in high-stakes clinical scenarios, machine learning models should provide well-calibrated measures of uncertainty alongside their predictions^7^. This allows identification of predictions that may be unreliable or unsafe if acted upon, and can help facilitate a “human-in-the-loop” system where decisions are triaged to a human healthcare worker if the model is sufficiently uncertain. Many modern machine learning models, such as deep neural networks, are too complex to be accurately assessed using traditional measures of uncertainty, such as confidence and prediction intervals, which are based on distributional or asymptotic assumptions that do not hold for most machine learning models. However, conformal prediction is a framework that can provide reliable uncertainty quantification for blackbox machine learning models^8,9^. Uncertainty quantification using conformal methods has received significant attention in the literature, and found success in applications to natural language processing ^10,11^, computer vision^12–14^, and biology^15,16^. The prediction intervals or sets from a conformal prediction procedure have exact, finite sample guarantees, e.g. an 95% prediction interval will cover the true value at least 95% of the time, regardless of the underlying data distribution or model used. Standard conformal prediction only requires exchangeability between the training and evaluation data distributions, which is a much weaker assumption than the independence assumptions needed by classical methods. However, the exchangeability assumption is often violated in real-world settings, but techniques like weighted conformal inference^17^ can be used to retain the desired coverage even in the presence of exchangeability violations. A common violation arises from so-called *covariate shift*^5,18^ wherein the marginal distribution of the inputs changes between different settings, but the conditional distribution that relates the inputs the outputs is invariant. This situation is common in many healthcare applications where the demographics of patient populations can vary between different hospitals, but the risk for two individuals with the same covariates remains the same across the different populations.

In this work, we demonstrate how weighted conformal prediction can enable the reliable transfer of prediction models between healthcare settings without collecting new labeled data. Given only covariate data from a new clinical environment, weighted conformal inference provides prediction sets that reflect expected changes in model performance due to covariate shift. This allows practitioners to determine when model predictions may be safely used in a new setting versus when additional verification is required. By leveraging distributional information from covariates alone, weighted conformal prediction facilitates model transport, overcoming the constraints posed by data privacy protections and the expense of gathering new labels. Our approach promises to make machine learning more accessible for sensitive and critical applications like healthcare.

## Results

### Cohort Description

First, we investigate whether there are significant shifts in patient covariate distributions between real hospitals – a phenomenon known as *covariate shift*. We used the eICU Collaborative Research Database^19^ to define mortality, phenotype, and remaining length of stay prediction (RLOS) tasks (**Figure 1**). Our base cohort comprised 73,718 unique patients with 20 shortlisted clinical variables. Detailed information regarding patient cohort and the shortlisted features for each admission can be found in Table 1 and Table 2, respectively. In total, we selected 16 hospitals for the mortality prediction task, 7 hospitals for the phenotype prediction task, and 15 hospitals for the RLOS prediction task. Our selection criteria aimed at maximizing the number of unique hospital pairs while ensuring a sufficient number of patients per hospital to support reliable conclusions.

**Figure 1:**
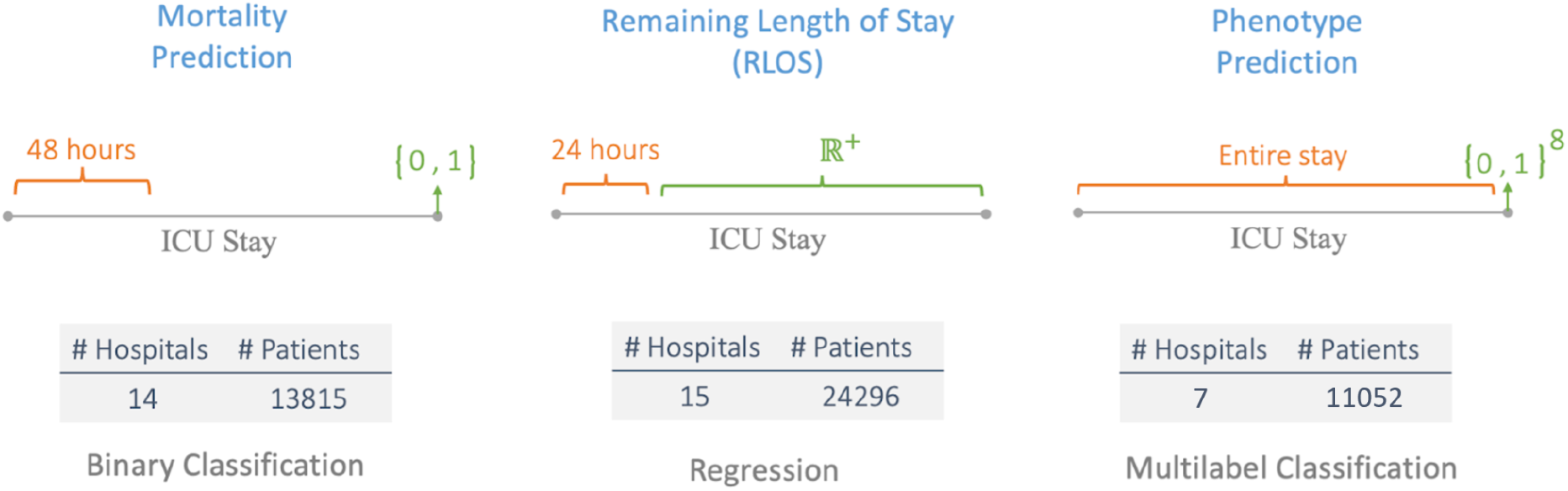
An array of tasks across different data modalities were chosen to evaluate the usefulness of the proposed workflow. Mortality prediction involved predicting a binary label for mortality using data from the first 48 hours of a patient’s ICU stay. A regression label was predicted for the patient’s remaining length of stay in the ICU using data from the first 24 hours. Phenotype prediction was defined as a multilabel prediction problem with 8 phenotypes using data from the entire ICU stay of a patient.

**Table 1:** Demographic and clinical profiles of ICU admissions, categorized by hospital outcomes. Continuous variables are presented as medians with interquartile ranges, while categorical variables are shown with their respective percentages. This table is reproduced following the eICU benchmarking paper^20^.

|  | Overall | Dead at Hospital | Alive at Hospital |
| --- | --- | --- | --- |
| **ICU Admissions** | 73,718 | 6,167 | 67,551 |
| **Age** | 62.41 [52-75] | 68.12 [59-80] | 61.8 [52-75] |
| **Gender (F)** | 33,544 (45.5) | 2,830 (45.8) | 30,714 (45.4) |
| *Ethnicity* |  |  |  |
| **Caucasian** | 56,973 (77.2) | 4,866 (78.9) | 52,107 (77.1) |
| **African American** | 7,982 (10.8) | 582 (9.4) | 7,400 (10.9 ) |
| **Hispanic** | 2,937 (3.98) | 226 (3.6) | 2,711 (4) |
| **Asian** | 1,174 (1.59) | 97 (1.5) | 1,077 (1.5) |
| **Native American** | 413 (0.56) | 42 (0.68) | 371 (0.54) |
| **Unknown** | 4,239 (5.7) | 354 (5.7) | 3,885 (5.7) |
| *Outcomes* |  |  |  |
| **Hospital length of stay (days)** | 5.29 [2.53-6.84] | 3.9 [1.42-5.22] | 5.41 [2.65-6.92] |
| **ICU length of stay (days)** | 2.32 [1.01-2.91] | 3.17 [1.19-4.43] | 2.24 [1-2.83] |
| **Hospital Death** | 6,167 (8.36) | 6,167 (100) | - |
| **ICU Death** | 4,575 (6.2) | 4,575 (74.1) | - |

**Table 2:** List of clinical variables utilized in feature extraction and training of machine learning models. Variables are classified by data type, either numerical or categorical. This table is reproduced following the eICU benchmarking paper^20^.

| Variable | Data Type |
| --- | --- |
| Heart rate | Numerical |
| Mean arterial pressure | Numerical |
| Diastolic blood pressure | Numerical |
| Systolic blood pressure | Numerical |
| O2 | Numerical |
| Respiratory rate | Numerical |
| Temperature | Numerical |
| Glucose | Numerical |
| FiO2 | Numerical |
| pH | Numerical |
| Height | Numerical |
| Weight | Numerical |
| Age | Numerical |
| Admission diagnosis | Categorical |
| Ethnicity | Categorical |
| Gender | Categorical |
| Glasgow Coma Score Total | Categorical |
| Glasgow Coma Score Eyes | Categorical |
| Glasgow Coma Score Motor | Categorical |
| Glasgow Coma Score Verbal | Categorical |

### Assessment of Covariate Shift Between Hospitals

We used machine learning to quantify the extent of covariate shift among hospitals (see Methods). If patient demographics are consistent across two hospitals, a model should perform no better than random guessing in terms of classifying which hospital a given patient belongs to. If the demographics vary, the model should be able to correctly identify the hospital source. Our results indicate the strong presence of covariate shift between hospitals. As presented in Figure 2, the high AUROC (mean=0.94, SD=0.046) scores across tasks demonstrate that our models were consistently able to distinguish between hospitals. This discriminative ability signifies the existence of significant differences in the covariate distributions of various hospitals.

**Figure 2:**
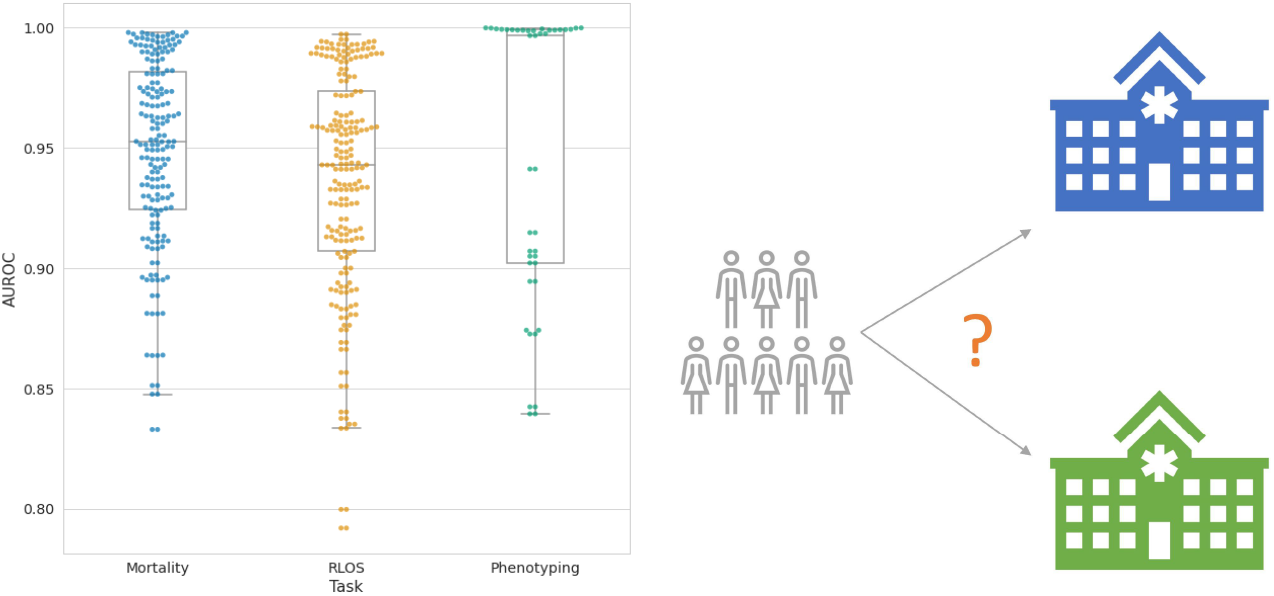
Covariate shift between hospitals was assessed by computing the area under the receiver operating characteristic curve (AUROC) score for a random forest classifier trained to distinguish between hospitals based on their patient covariates. A high score at this task suggests presence of patient covariate(s) that differ significantly from one hospital to the other.

### Evaluating the Impact of Covariate Shift on Model Performance

We investigated whether covariate shift affected the model’s predictive accuracy. **Figure 3A** depicts the performance of the models before and after transfer for each unique pair of hospitals. The majority of evaluations for mortality (87%) and phenotype (98%) tasks showed a decrease in performance after transfer, while the RLOS task (58%) showed a more mixed pattern. Across tasks, the median target hospital performance was consistently lower than the median source hospital performance (**Figure 3B**). These results underscore that covariate shift between hospitals predominantly leads to a decrease in the performance of transferred models.

**Figure 3:**
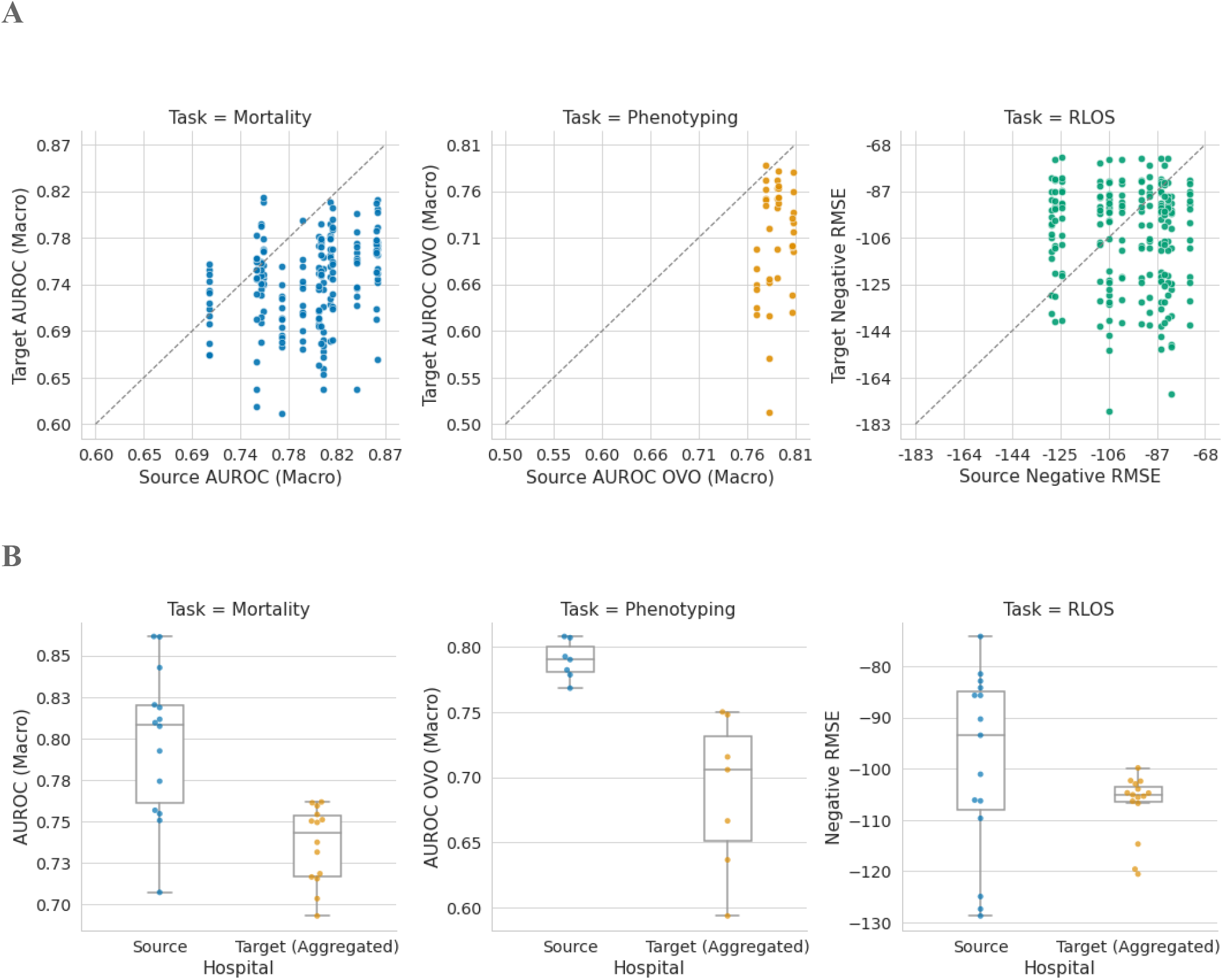
Covariate shift has a significant impact on the transferred model’s performance across all tasks. **Panel A**: The cross validation performance of a model on the source hospital data is compared with its performance on the target hospital data for each task. The diagonal line represents the case where covariate shift has no impact on model performance. **Panel B:** Aggregates the information from Panel A and contrasts cross validation performance on the source hospital data with average performance on the target hospital data for each task.

### Performance of Conformal Prediction in the Presence of Covariate Shift

We compared the marginal coverage and the average width of the prediction sets generated by the naive and the weighted conformal procedures for the transferred model (See Methods). If covariate shift accounts for the bulk of the observed distribution shift between the source and the target hospitals, we expect the weighted procedure to achieve marginal coverage greater than or equal to the chosen threshold of 90%, satisfying the coverage guarantee. The naive procedure may fail to satisfy the coverage guarantee as it assumes no distribution shift between source and target hospitals.

### Weighted conformal prediction obtains reliable uncertainty quantification in the presence of covariate shift

As depicted in **Figure 4A**, the estimated marginal coverage of the prediction sets, generated by both naive and weighted conformal procedures, varies significantly across each task and hospital pair. The naive procedure fails to meet the coverage guarantee in 63%, 78%, and 60% of evaluations across mortality, phenotyping, and RLOS tasks, respectively. In contrast, the weighted procedure demonstrated a much lower failure rate of 5%, 27%, and 10% for the same tasks, thereby supporting our hypothesis that covariate shift is the primary factor influencing distribution shift between source and target hospitals. Both conformal procedures exhibit their poorest performance on the phenotyping task, which we attribute to the task’s inherent complexity and the reduced size of the training datasets.

**Figure 4:**
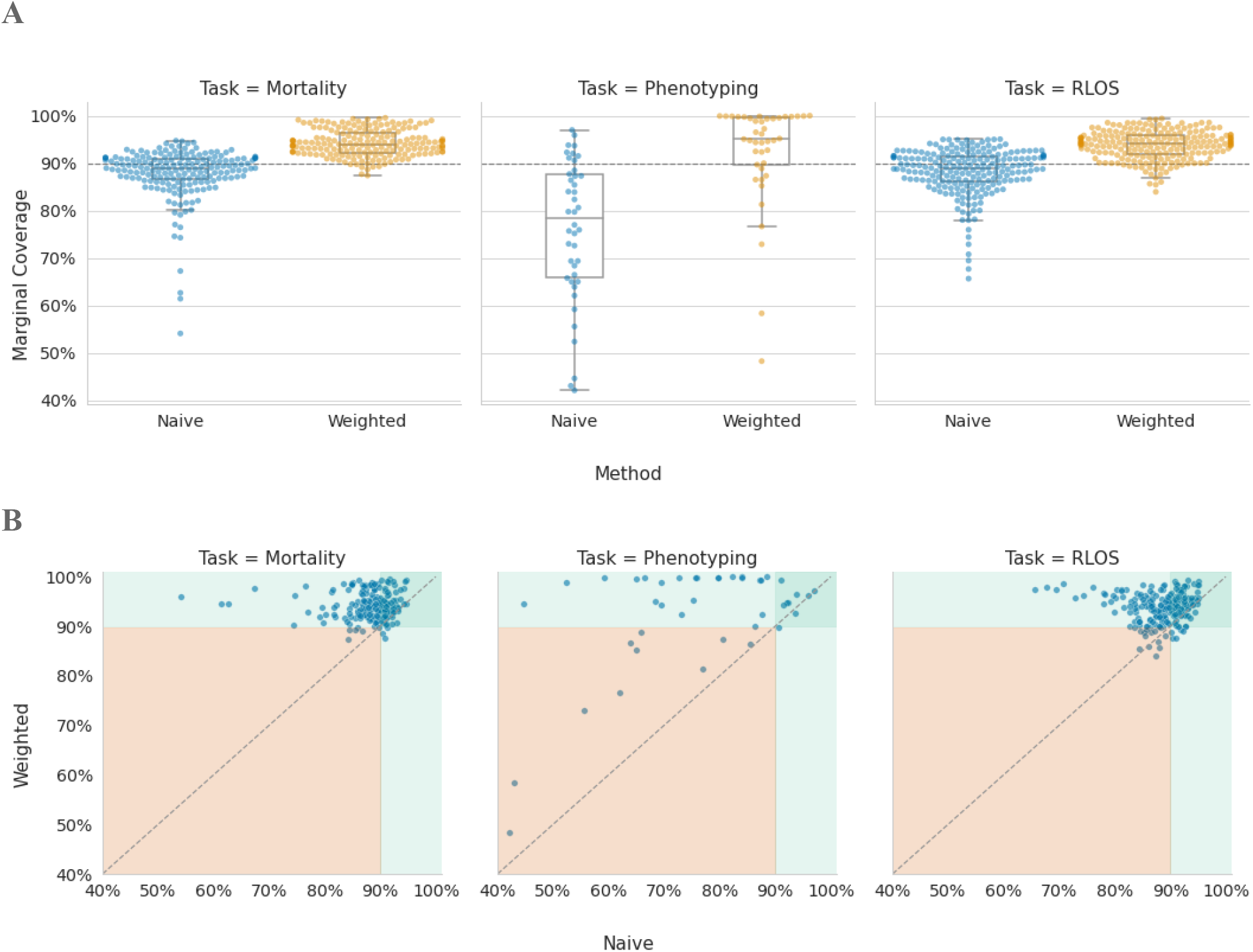
Covariate shift invalidates the theoretical guarantees of conformal prediction if used naively, resulting in prediction sets that have marginal coverage below the desired threshold of 90%. **Panel A**: A comparison between the naive and weighted conformal prediction methods across all 3 tasks and all hospital pairs. The naive approach fails to provide the correct coverage of 90%, while the weighted conformal method accounts for covariate shift to provide the correct level of coverage. **Panel B**: Difference in marginal coverage between the naive and weighted conformal methods across all 3 tasks for all hospital pairs. In general, the weighted approach results in a change in coverage that’s proportional to the difference between the desired threshold and naive prediction set coverage.

For a more granular comparison of the performance of the naive and weighted conformal procedures, we visualize the per-hospital-pair empirical coverage for the naive procedure against the weighted procedure for each task in **Figure 4B**. The weighted procedure showcases its capability to meet the coverage guarantee by adaptively improving over the coverage of the naive procedure. For instance, in mortality and RLOS tasks, the naive procedure can undercover by as much as 30% or as little as 1%. In both situations, the weighted procedure achieves coverage that meets the user desired threshold of 90%.

### Weighted conformal prediction sets are efficient

The simplest way to satisfy the coverage guarantee is to designate the entire set of possible labels as the prediction set. However, this approach isn’t efficient or useful. We investigate whether the satisfaction of coverage guarantee by the weighted conformal prediction is done in a trivial manner by evaluating the average width of the prediction sets generated by the naive and weighted conformal procedures. As shown in **Figure 5**, the average width of the prediction sets generated by the weighted procedure is larger than that of the naive procedure across tasks. Specifically, the weighted procedure had an average width of 1.64, 6.81, and 33.97, which was significantly larger than the naive procedure’s average width of 1.31, 4.85, and 7.9 for mortality, phenotyping, and RLOS tasks respectively. This is somewhat expected as it’s difficult to satisfy the coverage guarantee without expanding the prediction set while keeping the model fixed. However, the average width of the prediction sets generated by the weighted procedure is still smaller than the dimensionality of the label space in most cases, indicating a more efficient prediction. The phenotyping task is an exception, where the width of the prediction set often equals the dimensionality of the label space. However, even in these instances, the prediction set’s size can be informative for decision-making, as it may suggest the need to abstain from making a prediction when the prediction set is too wide.

**Figure 5:**
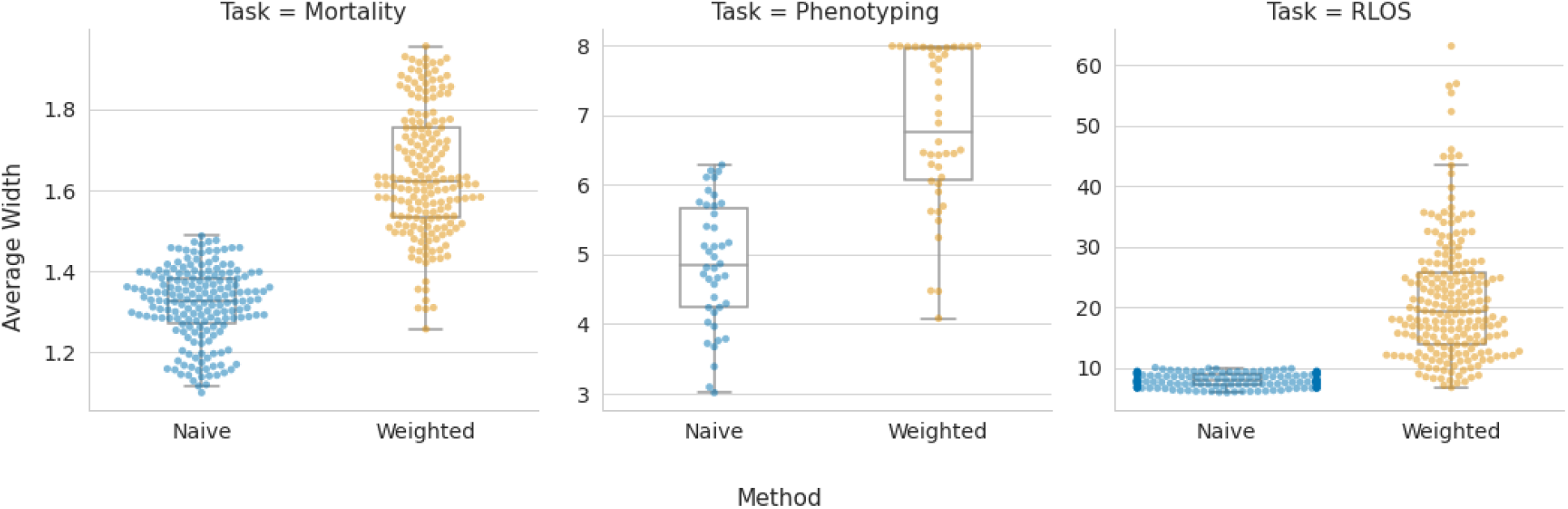
Assessment of prediction width. The naive conformal method was overly confident and produced prediction intervals that were too narrow to properly cover the true level at the correct rate. The weighted method maintains coverage while still producing meaningfully tight prediction sets.

### Fairness Assessment of Conformal Prediction Sets

While the weighted conformal procedure generally satisfies the marginal coverage guarantee across most hospital pairs and tasks, it is crucial to examine whether this guarantee is equitably satisfied across different patient populations. The marginal coverage, by its nature, averages performance over the entire test set, potentially obscuring disparities in performance across different patient strata. It is conceivable that the weighted conformal procedure might achieve the desired marginal coverage by satisfying the coverage guarantee for the majority patient strata, while failing to do so for minority strata. A similar pattern may also exist for the naive conformal procedure.

To investigate this, we present in Figure 6 the ethnicity-stratified marginal coverage for both the naive and weighted conformal procedures, across each hospital pair and task. While this analysis does not provide a comprehensive evaluation of fairness, it does offer some insight into the potential disparities. We observe that the naive conformal procedure satisfies the per-strata coverage guarantee for exactly 1 strata (out of 6) in the mortality prediction task, undercovering across other tasks and stratas. In contrast, the weighted procedure achieves the desired coverage for 5 stratas in mortality prediction task and 4 stratas in phenotype prediction task. For the RLOS task, although the weighted conformal procedure improved stratified marginal coverage compared to the naive conformal procedure, it did not achieve the desired coverage. It is important to note that the weighted conformal procedure is not designed to satisfy the coverage guarantee for all strata, as the weights are estimated to address covariate shift at the hospital level. Nevertheless, the ability of the weighted conformal procedure to satisfy the coverage guarantee for most strata across the mortality prediction and phenotyping tasks is promising.

**Figure 6:**
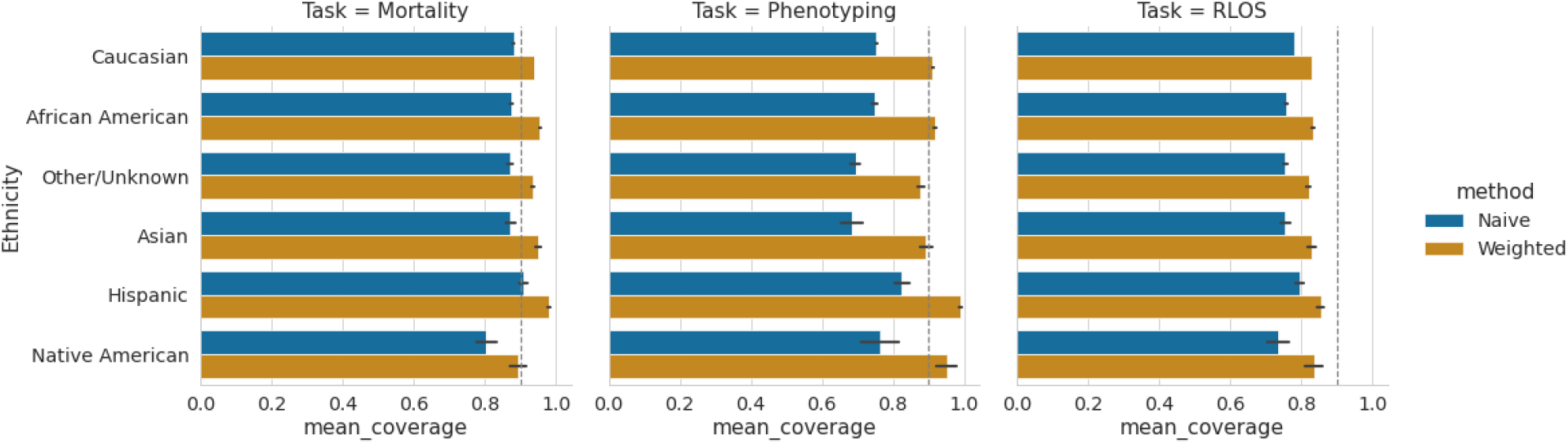
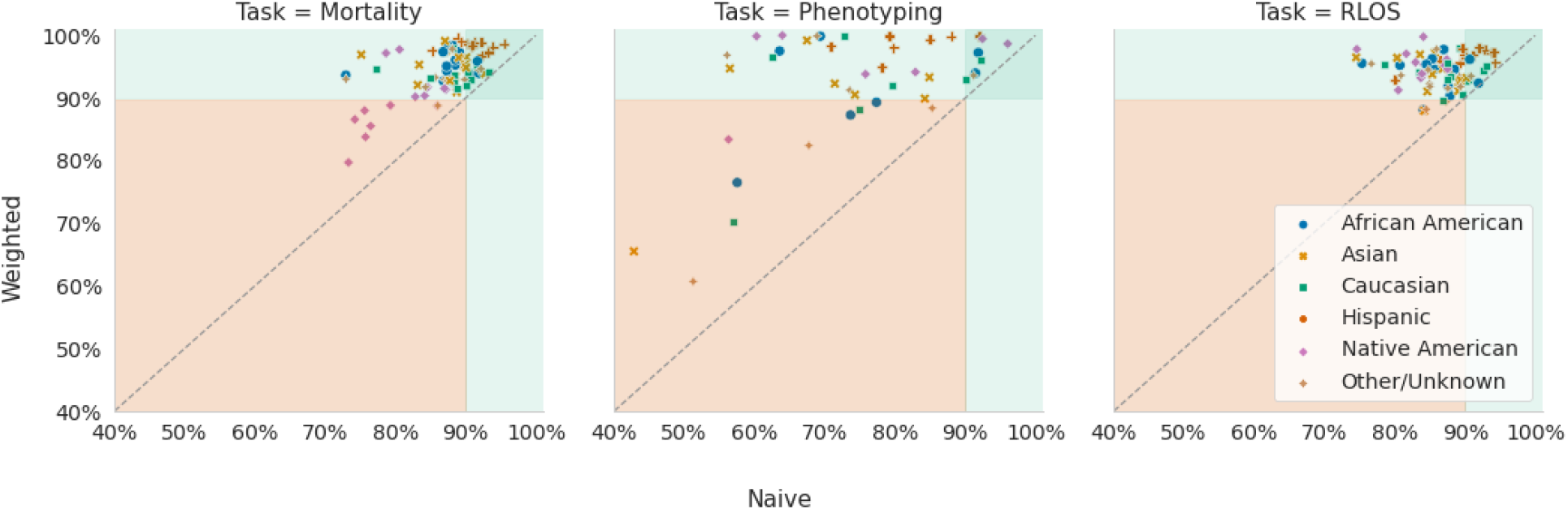
Naive conformal prediction sets were found to have marginal coverage below the desired threshold when the target hospital’s patient data was stratified based on ethnicity. Weighted conformal method was able to remedy the impact of covariate shift and resulted in prediction sets that satisfied the marginal coverage guarantee in almost all stratas for mortality prediction and phenotyping. For remaining length of stay prediction, the weighted conformal prediction sets had much higher marginal coverage than naive conformal prediction sets, however, not to the level that was desired.

## Discussion

Our study presents a novel approach to addressing a key challenge in healthcare: the reliable transfer of machine learning models across different clinical settings. We demonstrated that weighted conformal prediction effectively manages covariate shift, a prevalent issue when deploying models in new healthcare environments. This methodology provides a valuable tool for clinicians and practitioners, offering a practical solution for model transfer without necessitating the collection of new labeled data. The presence of covariate shift, as evidenced by our analysis, significantly impacts the performance of predictive models when transferred between hospitals. Traditional methods of adapting models, such as retraining or fine-tuning with local data, are often constrained by resources and data availability. Our approach circumvents these limitations by utilizing weighted conformal prediction, which leverages existing covariate data to adjust for distributional changes, thereby ensuring reliable model performance.

Our findings indicate that weighted conformal prediction in healthcare can facilitate the transfer of machine learning models across clinical settings without the need for new labeled data, broadening their accessibility and use. This approach not only streamlines model adaptation, but also significantly reduces resource expenditure, a key advantage for resource-constrained healthcare environments. However, ensuring equitable performance and addressing potential biases in predictions across various patient groups is an area that requires further research to ensure the fair and ethical application of these tools.

Future research should focus on extending this methodology to a wider range of clinical tasks and exploring its applicability in other domains where covariate shift is a concern. Additionally, a deeper exploration into the ethical implications and fairness of the model’s predictions across diverse patient populations is essential to ensure equitable healthcare outcomes.

In conclusion, our study highlights the potential of weighted conformal prediction in overcoming the challenges of model transfer in healthcare. This approach represents a significant step forward in making advanced predictive tools more accessible and reliable across various clinical settings.

## Limitations

Our study’s limitations stem primarily from the context-specific nature of our findings. The results apply specifically to the scenario of covariate shift and may not address other prevalent shifts like concept drift or label shift. Potentially, the poor performance on the phenotyping task can be attributed to such shifts. Moreover, the quality and diversity of the source data are critical; biases in the training data could be perpetuated despite adjustments made by our method. Further, the improved uncertainty quantification does not necessarily equate to improved model accuracy. This aspect is particularly vital in high-stakes scenarios such as healthcare, where ethical considerations and potential biases in model predictions need careful examination. Additionally, the use of retrospective data in our research, while comprehensive, may not entirely reflect the complexities of real-time clinical settings.

## Data Availability

All data produced in the present study are available upon reasonable request to the authors

## Author contributions

### Acknowledgement

The authors wish to disclose that ChatGPT based on GPT-4 was used to edit and revise some passages in this paper. Specifically, ChatGPT was used as a writing aid to assist in refining human-generated text to enhance the overall clarity of the writing. However, the intellectual contributions, ideas, and conclusions presented herein are exclusively the product of the authors.

### Funding Sources

ALB was partially supported by an award from the National Heart, Lunch, and Blood Institute (HL141771).

## Materials and Methods

### Data Preprocessing and Cohort Assembly

Our study utilized the eICU Collaborative Research Database^19^. The eICU database is a multi-center intensive care database comprising 200,859 admissions to 208 hospitals in the United States between 2014 and 2015. Following the inclusion criteria outlined in Sheikhalishahi et al.^20^, our base cohort comprised 73,718 unique adult (age > 18) patients, each with an ICU admission that had at least 15 records (Table 1). In relation to each patient admission, 20 features from the **patient** (administrative data and demographic details), **lab** (routine care laboratory measurements), **nurse charting** (bedside records), and **diagnosis** tables were shortlisted (Table 2). To mitigate data sparsity arising from irregular measurement intervals, we aggregated clinical variables into hourly intervals. Using the *tsfresh*^21^ Python package, we extracted features and imputed missing values, generating an aggregate covariate matrix with 306 time-series features for each patient

### Task definition

**Mortality prediction** was formulated as a binary classification task, predicting patient mortality using the first 48 hours of a patient’s ICU stay. We selected 16 hospitals for this task, each with more than 600 admissions. **Phenotype prediction** was characterized as a multilabel classification task, involving the prediction of 8 distinct phenotypes based on a patient’s entire ICU stay. The chosen phenotypes were prevalent across numerous hospitals, ensuring adequate patient representation per phenotype and hospital to produce meaningful results. In total, 7 hospitals were selected for this task, each with more than 100 admissions. Finally, **remaining length of stay prediction (RLOS) prediction** was defined as a regression task, estimating the number of days a patient would remain in the ICU based on their first 24 hours of ICU stay. For this task, we selected 15 hospitals, each with more than 1000 admissions.

### Machine learning models

We used two primary models for the classification tasks: logistic regression and random forest classifier. Both were chosen due to their resilience in handling expansive feature spaces and their limited need for hyperparameter tuning. For the RLOS task, a penalized linear regression model and a random forest regressor were utilized. All the models were implemented using the scikit-learn Python package^22^.

### Nested Cross Validation

To ensure the validity and generalizability of our results, we employed a nested cross-validation strategy. It consists of two nested loops. Within the inner cross-validation loop, various models are trained and evaluated on subsets of the data to identify the best-performing model for each hospital. This loop primarily handles model selection. Once the best model for each hospital has been identified in the inner loop, the outer cross-validation loop tests these models on unseen data. This helps in removing the optimistic bias that arises from using the same data for both model selection and evaluation.

### Evaluation Metrics

We employed the Area Under the Receiver Operating Characteristic Curve (AUROC) metric for evaluating model performance in binary classification tasks, such as covariate shift assessment and mortality prediction. AUROC assesses the discriminative performance of a binary classifier and is insensitive to class imbalance and the choice of decision threshold. An AUROC of 1 signifies a perfect classifier, while an AUROC of 0.5 indicates a classifier performing at the level of random guessing. The phenotyping task was evaluated using the macro-weighted AUROC, which is the simple average of the AUROC obtained in each component binary classification task

For the Remaining Length of Stay (RLOS) prediction task, we utilized the negative root mean squared error (RMSE) metric to evaluate model performance. This metric provides a measure of the deviation between the predicted and actual values, with lower values indicating superior performance.

### Density ratio estimation

The density ratios between patient covariates from a source hospital and a target hospital were estimated by fitting a probabilistic binary classifier. This classifier was designed to predict whether a patient’s covariates originated from the source hospital or the target hospital.

Specifically, let 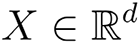 represent the patient covariates and 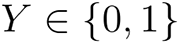 denote the label, where *Y* = 1 indicates that the patient covariate is from the target hospital and 0 otherwise. A probabilistic classifier 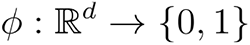 was trained to estimate the conditional density of *Y* given *X*. The density ratio between the target and the source hospital for a given patient covariate *x* was obtained by substituting estimates 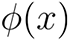 for 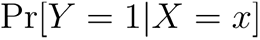 and 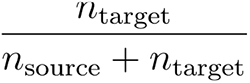 for 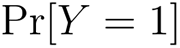 in the following:

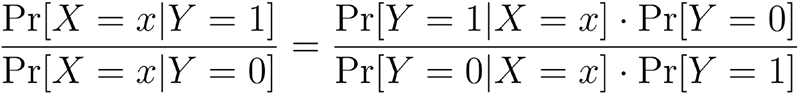

### Covariate Shift Assessment

We estimated the performance of a binary classifier in distinguishing between the source and target hospitals using nested cross validation. This setup mirrored the process used in density ratio estimation. The AUROC obtained by the classifier was employed as the covariate shift metric. Higher AUROC values indicate the classifier’s reliable differentiation between source and target hospitals, thereby suggesting the presence of a covariate shift

To further investigate the impact of covariate shift on model performance, we first obtained an unbiased estimate of the performance of each model on the source hospital data using nested cross-validation. Then, we trained the optimal model configuration on the entire dataset of the source hospital and evaluated its performance on the target hospital data. This process was repeated for each unique pair of hospitals. For the RLOS task, we used the negative RMSE as the performance metric, while for the mortality and phenotype tasks, we employed the macro-weighted AUROC.

### Prediction Sets

A prediction set for a test instance 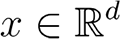 is a subset of the label space *Y*. The size of the prediction set indicates model confidence, with larger prediction sets indicating greater uncertainty.

### Marginal Coverage

The expected proportion of test prediction sets that contain the true label is referred to as the marginal coverage. It is a measure of reliability of uncertainty quantification algorithms. When the marginal coverage for an algorithm is greater than or equal to a user-specified threshold, the algorithm is said to satisfy the marginal coverage guarantee.

### Average Width

The average size of prediction set for a test instance is referred to as the average width. It offers a measure of efficiency of uncertainty quantification algorithms. When comparing two algorithms with equal reliability, the one with the smaller average width is typically more desirable.

### Standard Conformal Prediction

Conformal prediction is a model-agnostic method for uncertainty quantification that necessitates minimal data distribution assumptions to provide theoretical reliability guarantees. It wraps any trained machine learning model and a labeled calibration dataset to create prediction sets for test instances that satisfy the user specified marginal coverage guarantee. Critical to the reliability of conformal prediction is the assumption that the training, calibration, and test data are drawn from the same distribution exchangeably. This assumption is violated in the presence of covariate shift, as the source (train, calibration) and target (test) hospitals have distinct covariate distributions. Correspondingly, we refer to standard conformal prediction as naive conformal prediction in the text to underscore this violation.

The choice of nonconformity measure determines the efficiency of the conformal prediction procedure. We used the adaptive classifier^23^ nonconformity measure for the mortality prediction task. For phenotype prediction, which is a multilabel classification task, we derived a novel extension of the procedure (see Supplementary Materials). For the RLOS prediction task, we used the mean absolute error as the nonconformity measure.

### Weighted Conformal Prediction Procedure

To address covariate shift, we implemented a variant of conformal prediction that maintains theoretical reliability while being robust to covariate shift^17^. Since this method involves data reweighting, we refer to it in the text as weighted conformal prediction. The weighted method requires three components: unlabeled data from the target hospital, a labeled calibration dataset, and a trained machine learning model. The calibration and the model training data are assumed to be exchangeable. The unlabeled calibration and target hospital data are utilized to quantify the distribution shift between the source hospital and the target hospital. These estimates are subsequently used to derive weights that compensate for covariate shift and retain marginal coverage guarantee.

### Assessment of Conformal Prediction Methods

We implemented a 70%-30% train-calibration split of the source hospital data for both naive and weighted conformal prediction procedures. Given that the coverage guarantee is not conditional on a specific train-calibration split, we conducted our evaluation 100 times with random train-calibration splits. This approach allowed us to obtain a bootstrap estimate of the marginal coverage and average width. The density ratio estimator required by the weighted conformal procedure to account for any distribution shifts was trained on unlabeled data from both the source and target hospitals.

## Acknowledgement

The authors wish to thank Ben Kompa, Rachel Nethery, and Kevin Chen for their valuable feedback on an early version of this manuscript, and Amita Varma for her assistance in reviewing the final version of the write-up.

## Appendix

**Supplementary Figure.**
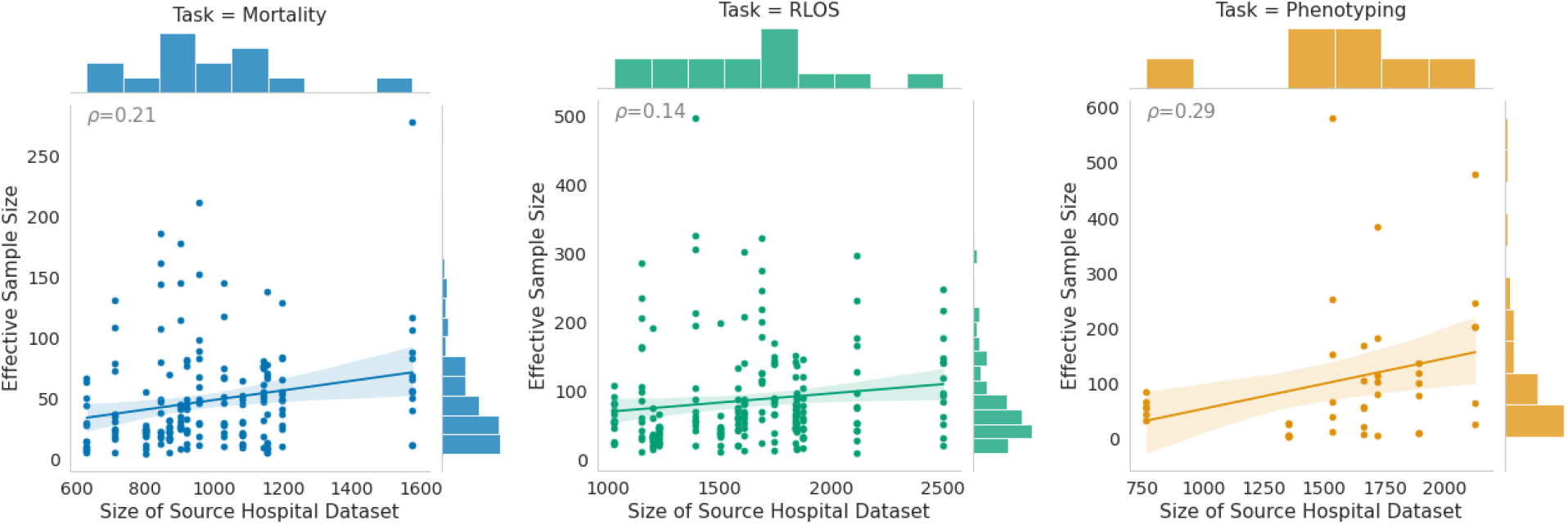
Importance weights were used to calculate the effective sample size of a source hospital’s dataset for a corresponding target hospital. A low effective sample size for a target hospital suggests that notwithstanding the size of a source hospital’s dataset, a machine learning model trained on the source hospital’s dataset is unlikely to generalize to target hospital’s dataset.

## References

1. Beam, A. L. & Kohane, I. S. Big data and machine learning in health care. JAMA – Journal of the American Medical Association vol. 319 1317–1318 Preprint at 10.1001/jama.2017.18391 (2018).

2. Finlayson, S. G., Beam, A. L. & van Smeden, M. Machine Learning and Statistics in Clinical Research Articles—Moving Past the False Dichotomy. JAMA Pediatr. (2023).

3. Beam, K., Sharma, P., Levy, P. & Beam, A. L. Artificial intelligence in the neonatal intensive care unit: the time is now. J. Perinatol. (2023) doi:10.1038/s41372-023-01719-z.

4. Wong, A. et al. External validation of a widely implemented proprietary sepsis prediction model in hospitalized patients. JAMA Intern. Med. 181, 1065–1070 (2021).

5. Finlayson, S. G. et al. The clinician and dataset shift in artificial intelligence. N. Engl. J. Med. 385, 283–286 (2021).

6. McKinney, S. M. et al. International evaluation of an AI system for breast cancer screening. Nature 577, 89–94 (2020).

7. Kompa, B., Snoek, J. & Beam, A. L. Second opinion needed: communicating uncertainty in medical machine learning. npj Digital Medicine 4, 1–6 (2021).

8. Shafer, G. & Vovk, V. A tutorial on conformal prediction. arXiv [cs.LG] (2007).

9. Angelopoulos; Stephen Bates., A., Angelopoulos, A. N. & Stephen Bates (Computer scientist). Conformal Prediction: A Gentle Introduction. (Now Publishers, 2023).

10. Quach, V., et al. Conformal Language Modeling. *arXiv [cs.CL]* (2023).

11. Kumar, B., et al. Conformal Prediction with Large Language Models for Multi-Choice Question Answering. in ICML workshop on Trustworthy, Enhanced, Adaptable, Capable and Human-centric (TEACH). arXiv preprint arXiv:2305.18404 (2023).

12. Kumar, B., Palepu, A., Tuwani, R. & Beam, A. Towards Reliable Zero Shot Classification in Self-Supervised Models with Conformal Prediction. in NeurIPS Workshop on Self-Supervised Learning (2022).

13. Wieslander, H. et al. Deep Learning With Conformal Prediction for Hierarchical Analysis of Large-Scale Whole-Slide Tissue Images. IEEE J Biomed Health Inform 25, 371–380 (2021).

14. Angelopoulos, A., Bates, S., Malik, J. & Jordan, M. I. Uncertainty Sets for Image Classifiers using Conformal Prediction. arXiv [cs.CV] (2020).

15. Fisch, A., Schuster, T., Jaakkola, T. & Barzilay, D. R. Conformal Prediction Sets with Limited False Positives. in Proceedings of the 39th International Conference on Machine Learning (eds. Chaudhuri, K. et al.) vol. 162 6514–6532 (PMLR, 17--23 Jul 2022).

16. Fannjiang, C., Bates, S., Angelopoulos, A. N., Listgarten, J. & Jordan, M. I. Conformal prediction under feedback covariate shift for biomolecular design. Proc. Natl. Acad. Sci. U. S. A. 119, e2204569119 (2022).

17. Tibshirani, R. J., Foygel Barber, R., Candes, E. & Ramdas, A. Conformal prediction under covariate shift. Adv. Neural Inf. Process. Syst. 32, (2019).

18. Failing loudly: An empirical study of methods for detecting dataset shift. https://proceedings.neurips.cc/paper/2019/hash/846c260d715e5b854ffad5f70a516c88-Abstract.html.

19. Pollard, T. J. et al. The eICU Collaborative Research Database, a freely available multi-center database for critical care research. Sci Data 5, 180178 (2018).

20. Sheikhalishahi, S., Balaraman, V. & Osmani, V. Benchmarking machine learning models on multi-centre eICU critical care dataset. PLoS One 15, e0235424 (2020).

21. Maximilian Christ, Nils Braun, Julius Neuffer, Andreas W. Kempa-Liehr. Time Series FeatuRe Extraction on basis of Scalable Hypothesis tests (tsfresh – A Python package). Neurocomputing 307, 72–77 (2018).

22. Pedregosa, F. et al. Scikit-learn: Machine Learning in Python. arXiv [cs.LG] 2825–2830 (2012).

23. Romano, Y., Sesia, M. & Candes, E. Classification with Valid and Adaptive Coverage. Adv. Neural Inf. Process. Syst. 33, 3581–3591 (2020).

